# Remote blood sampling differential protein expression associated with persistent hypertension following a hypertensive disorder of pregnancy

**DOI:** 10.1101/2025.07.30.25332389

**Authors:** Brian Kleiboeker, Janet Catov, Koen Raedschelders, Aleksandra Binek, Simion Kreimer, Noel Bairey Merz, Jennifer Van Eyk, Arun Jeyabalan, Adam C. Straub, Agnes Koczo, Alisse Hauspurg

## Abstract

**Objective:** Hypertensive disorders of pregnancy are associated with future cardiovascular risk, however, the underlying mechanisms are unclear. We sought to test the feasibility of postpartum remote blood sampling following a hypertensive disorder of pregnancy and subsequently identify differentially expressed proteins in individuals who developed hypertension.

**Study design:** We used data from a randomized clinical trial evaluating the feasibility of lifestyle intervention and home blood pressure monitoring of individuals with pre-pregnancy body mass index ≥25 kg/m^2^ and new-onset hypertensive disorders of pregnancy. Blood pressure was measured at remote visits at 6 weeks and 1 year postpartum and blood microsamples were collected at the second remote visit. Mass spectrometry was used to quantify 381 proteins, from which differential expression and pathway enrichment analyses were performed to detect alterations with persistent hypertension.

**Results:** Of the 100 randomized individuals, 87 completed sample collection with 85 yielding usable proteomics data. 4 proteins were differentially expressed with false discovery rate < 0.05 in individuals not taking antihypertensive medications who developed Stage 2 hypertension: Complement C4-B (C4B) and inter-alpha-trypsin inhibitor heavy chain 4 (ITIH4) levels were elevated while Afamin (AFM) and myosin light chain 6 (MYL6) levels were decreased. Pathway enrichment analysis suggests a shared function of inhibition of endopeptidases (such as Neprilysin, which degrades natriuretic peptides) in upregulated proteins and ubiquitin-proteasome mediated proteolysis activity in downregulated proteins.

**Conclusions:** Home microsampling is a promising methodology for postpartum sample collection. Serum proteomics using this approach suggest that protein homeostasis may be dysregulated in persistent hypertension following hypertensive disorders of pregnancy, providing novel candidates for future interventions.

**Condensation page:** **1) Tweetable statement: a short condensation of the paper, consisting of no more than 210 characters, stating its essential point(s).** Proteomic analysis reveals that de novo hypertension after hypertensive disorders of pregnancy correlates with dysregulated protein homeostasis.

**3) AJOG at a Glance: This section only applies to Original Research and Systematic Review submissions. This section is limited to no more than 130 words, 1-3 short sentences or phrases in bullet form, briefly describing your study, its significance, and its contribution to the literature. Authors should minimize the use of abbreviations (only very commonly used abbreviations will be allowed) and define an abbreviation prior to use in this section. Responses should be listed in bullet form after the A., B., and C. headings as below (not in paragraph form). All responses are subject to minor editorial alterations and/or shortened without the authors’ approval, and published both in print and on the Journal website**.

*Why was this study conducted?:*

- Prioritization of newborn care during the early postpartum months makes in-person research study visits challenging, necessitating improved methodologies.
- Persistent postpartum hypertension may mediate the established relationship between hypertensive disorders of pregnancy and future cardiovascular disease risk, yet its pathogenesis remains poorly understood.

*What are the key findings?:*

- Home microsampling is a promising methodology for postpartum sample collection.
- Serum proteomics using this approach suggest that protein homeostasis may be dysregulated in persistent hypertension following hypertensive disorders of pregnancy.

*What does this study add to what is already known?:*

- Our findings suggest that certain known features of hypertensive disorders of pregnancy, such as coagulation cascade activation, alterations of serum Afamin and proteasome levels, and protease dysregulation, might persist and contribute to de novo postpartum hypertension.
- Some of these pathways may be amenable to future intervention development to prevent persistent hypertension after a hypertensive disorder of pregnancy.

## INTRODUCTION

Hypertensive disorders of pregnancy (HDP) are associated with future cardiovascular risk in multiple, diverse cohorts. The biologic pathways linking HDP to cardiovascular disease (CVD) are not well characterized. Prior studies have shown a 40-50% risk of development of hypertension in the first year after delivery following severe preeclampsia.^1^ Overweight and obese individuals with HDP are at even higher risk.^2–4^ At the time of diagnosis and in the immediate postpartum period, those with a HDP have significant cardiovascular dysfunction with endothelial disruption, coagulation cascade activation and systemic inflammation.^5^ Whether these changes persist and are underlying drivers of development of hypertension or long-term CVD is unclear. Understanding biological pathways linking HDP to development of hypertension may yield new targets for intervention in the peri– and postpartum period to reduce risk of CVD and future pregnancy complications.

One challenge to studying individuals in the postpartum period is retention and attendance at in-person research study visits, given the prioritization of newborn care during the early postpartum months.^6^ In an effort to continue research without in-person contact during the COVID-19 pandemic, many studies developed protocols for remote research study visits.^7^ Development of low blood volume microsampling devices allows for collection of several microliters of blood without an in-person research study visit. Such remote microsampling protocols might also have utility in postpartum research.

In this study, we tested the feasibility of remote blood sample collection at a postpartum research study visit among a cohort with HDP. Furthermore, we sought to use this remote blood sampling methodology to identify differential protein biomarker expression among overweight and obese individuals with de novo HDP (without chronic hypertension) who progressed to hypertension in the first year postpartum compared to those who remained normotensive.

## MATERIALS AND METHODS

### Description of parent study

Heart Health 4 New Moms (HH4NM) was a pilot, prospective randomized trial (NCT03749746) evaluating the feasibility and impact of a lifestyle intervention and home blood pressure monitoring in the first year after delivery in individuals with overweight and obesity (pre-pregnancy body mass index [BMI] ≥25 kg/m^2^) and HDP. Full details of the parent study have been previously published.^3^ Briefly, eligible individuals had no pre-pregnancy hypertension and had a singleton pregnancy complicated by a HDP (gestational HTN, preeclampsia without severe features, or preeclampsia with severe features) and delivered between January 2019 and May 2021. Recruitment occurred postpartum across two sites in one state and randomization was 1:1:1 to usual care, home BP monitoring program alone, or home BP monitoring plus an internet-based lifestyle program.

All participants underwent assessment at two timepoints: baseline (6 weeks to 6 months postpartum) and at study completion (8-12 months postpartum) after at least 6 months in the study. HDP diagnoses were adjudicated by physicians (AH and AJ) using criteria from the American College of Obstetricians and Gynecologists (ACOG). At study visits, blood pressure was measured at rest, after sitting for at least 5 minutes by study staff (pre-pandemic) or self-collected with study staff observing and recording results using a secured Zoom room.^3^ Blood pressure was measured in triplicate using either an iHealth, Wireless Blood Pressure Monitor BP5 or an A&D UA-651 (A&D Medical; San Jose, California) automatic upper arm blood pressure monitor, validated by Dabl Educational Trust and the British Society for Hypertension for use in pregnant and postpartum individuals. The mean of the three measures was used in analysis. The primary outcome of the trial was feasibility. No differences were seen in weight or blood pressure between the three study arms. As such, randomization groups were combined for this analysis.

### Home blood sample collection and processing

At the time of the second study visit (46.2 ± 8.0 weeks postpartum), individuals were invited to provide a home-collected blood sample using a Neoteryx Mitra clamshell device, which allows for self-collection of up to 30 uL of blood. A pre-paid envelope containing desiccant was provided to participants to mail the device back to study staff. Protein content was extracted from the Mitra devices by 1 hour incubation at 60°C in 35% TFE buffer containing 40 mM dithiothreitol (DTT; Sigma) and 50 mM ammonium bicarbonate (Sigma). The remainder of sample processing was performed automatically on a Beckman i7 automated workstation.

Iodoacetamide was added to 10 mM concentration and the samples were incubated at 25°C in the dark for 30 minutes. An addition of 5 mM DTT quenched alkylation and addition of 50DmM NH4CO3 diluted TFE to 5% final concentration. Trypsin was added at a ratio of 25:1 and samples were digested for 4 hours at 43°C. Digestion was quenched with addition of formic acid to 1%. Samples were diluted to 1000 ng/20 uL concentration before liquid chromatography and quantification by mass spectrometry (LC-MS) analysis.

### LC-MS analysis

Full details of the LC-MS analysis have been previously published.^8^ Briefly, samples were analyzed on an Ultimate 3000 nano-RSLC equipped with the capillary flow selector at 9.5 uL/min analytical flowrate. Peptides were trapped on Phenomenex Kinetex C8 HPLC column (150 mm length, 0.3 mm inner diameter, 5 um beads) and separated on a CapLC 50 cm micro-pillar array column from Pharmafluidics all at 50°C. Gradient conditions were: start at 7% B; 7-22% B over 8.5 minutes; 22-38% B over 4.3 minutes; jump to 98% B over 0.2 minutes; 98% B,

0.9 minutes; 7% B, 1 minute (mobile phase A, 0.1% formic acid in water; mobile phase B, 0.1% formic acid in acetonitrile). Loading conditions were: 50% B, 50% C, 60 uL/min down to 55 uL/min over 0.5 minutes; switch to 100% A over 0.3 minutes; 100% A for 8.2 minutes (loading buffer A, 0.1% formic acid in water; B, 0.2% formic acid in 70% acetonitrile 30% water with 5 mM ammonium formate; C, 0.2% formic acid in 90% isopropanol with 10% acetonitrile and 5 mM ammonium formate). The loading pump flowrate was reduced to 20 uL/min at 9.5 minutes and held for the remainder of the method for sample loading and desalting.

Data were acquired on a Bruker TIMS-TOF Pro mass spectrometer using an accumulation time of 70 ms, separation ramp of 70 ms, and fragmentation windows of 40 m/z which covered 360 to 1120 m/z and 0.65 to 1.41 1/K0 ion mobility ranges. Each ramp cycle utilized 1 to 3 DIA over a total cycle time of 0.76 seconds. Electrospray ionization was performed using a Bruker MnESI source with an 8 nozzle M3 emitter (Newomics) with a capillary voltage of 4800 V, endplate offset voltage of 500 V, the nebulizer set to 2.9 Bar, and the dry gas set to 6.0 L/min and 200°C. Raw data were analyzed using dia-PASEF.^9^

### Subgroup Classification of Postpartum Hypertension

All individuals who completed both study visits and provided a home blood sample were included in the analysis. Blood pressure categories were defined per American Heart Association guidelines.^10^ Briefly, normal blood pressure was defined as systolic blood pressure (SBP) <120 mmHg and diastolic blood pressure (DBP) <80 mmHg. Individuals with SBP 120-129 mmHg and normal DBP were considered to have elevated blood pressure. Stage 1 HTN was defined as SBP 130-139 mmHg or DBP 80-89 mmHg. Stage 2 HTN was defined as 1.) SBP ≥140mmHg or DBP ≥90mmHg or 2.) requiring treatment with antihypertensive medications. Individuals were classified as having persistent postpartum hypertension if they had at least Stage 1 HTN or greater.

Linear regression was performed to test for proteins differentially abundant in individuals with Stage 1 or Stage 2 HTN. For Stage 2 HTN, two comparisons were made: 1) including all individuals with Stage 2 HTN by the above definition and 2) following disaggregation by medication status to include only individuals not taking antihypertensive medications. The latter comparison was made to explore the relationship between uncontrolled Stage 2 HTN and serum protein levels with the aim of better understanding the pathogenesis of HTN in the postpartum period. Some effects of antihypertensives on plasma protein levels are known (e.g. ACE inhibitors, β-blockers, and α-methyldopa altering renin levels^11–13^), but it is likely that many others are unknown and could thus confound this aim.

Secondary analysis was performed with a separate classification which compared blood pressures between two visits. Based on these comparisons, participants were classified into one of three groups: improved HTN, worsened HTN, or no change in HTN. Participants were considered to have improved HTN if their blood pressure classification improved from the first to the second visit (Stage 2 to Stage 1 HTN, Stage 2 HTN to normotension, or Stage 1 HTN to normotension) or if the participant discontinued antihypertensive medications between the two visits with no change in blood pressure. Participants were classified as having worsened HTN if their blood pressure classification worsened between visits.

### Statistical analysis

This analysis relating serum biomarkers with persistent postpartum HTN was pre-specified, but the comparisons reported were chosen post-hoc. All statistical analyses and data visualization were performed in the R statistical programming language. Differences in BMI between groups were analyzed using the non-parametric Kruskal-Wallis rank sum test, while all other continuous and categorical variables were analyzed with a one-way analysis of means or Fisher’s exact test, respectively. For analysis of proteomic data, protein-level intensities from dia-PASEF were normalized using variance stabilization normalization.^14^ One outlier sample was removed which had only 46 proteins detected compared to 372.6 ± 6.6 (mean ± standard deviation) proteins detected for the other 85 samples. Although only 2.25% of the data were missing following removal of this outlier, GSimp was used to impute the missing values.^15^ **Table 1** was prepared using gtsummary.^16^ Pathway enrichment and differential expression analysis across blood pressure groups or blood pressure changes across visits was conducted using limma (linear regression) with BMI, tobacco smoking status, and self-reported race (as a social construct) as covariates.^17^ P-values were corrected for multiple comparisons using the

**Table 1:** Participant characteristics and obstetric outcomes of the 85 individuals included in this analysis. One-way analysis of means was used for age and gestational age at delivery, Kruskal-Wallis rank sum test was used for pre– and post-pregnancy BMI, and Fisher’s exact test was used for all categorical variables. HTN = hypertension, BMI = body mass index, HDP = hypertensive disorder of pregnancy, IQR = interquartile range.

| Characteristic | Normotensive,<br>N = 35 | Stage 1<br>hypertension, N<br>= 28 | Stage 2<br>hypertension, N<br>= 11 | Stage 2 HTN<br>(antihypertensive<br>medications), N = 11 | P-value<sup>†</sup> |
| --- | --- | --- | --- | --- | --- |
| Age (years), Median (IQR) | 32.5 (29.5 – 35.3) | 33.1 (29.7 – 35.2) | 34.8 (31.1 – 35.6) | 32.3 (31.1 – 36.5) | 0.57 |
| BMI (kg/m<sup>2</sup>), Median (IQR) | 31.5 (28.2 – 34.9) | 31.5 (29.4 – 36.4) | 39.0 (35.2 – 45.0) | 37.6 (30.5 – 42.8) | 0.006 |
| BMI pre-pregnancy (kg/m<sup>2</sup>), Median (IQR) | 30.3 (27.0 – 34.0) | 31.8 (28.9 – 34.8) | 33.4 (31.7 – 42.1) | 34.7 (30.7 – 36.7) | 0.029 |
| Race, n (%) |  |  |  |  | 0.002 |
| Black | 1 (2.9) | 10 (36) | 3 (27) | 2 (18) |  |
| Other | 0 (0) | 1 (3.6) | 1 (9.1) | 0 (0) |  |
| White | 34 (97) | 17 (61) | 7 (64) | 9 (82) |  |
| Reported smoking, n (%) | 5 (14) | 2 (7.1) | 1 (9.1) | 3 (27) | 0.39 |
| Gestational age at delivery (weeks), Median (IQR) | 38.1 (37.5 – 39.6) | 37.3 (36.1 – 37.6) | 37.4 (37.0 – 38.5) | 37.3 (34.7 – 38.6) | 0.031 |
| Preterm delivery (<34 weeks), n (%) | 1 (2.9) | 5 (18) | 2 (18) | 1 (9.1) | 0.14 |
| Gestational diabetes, n (%) | 3 (8.6) | 2 (7.1) | 1 (9.1) | 2 (18) | 0.82 |
| Type of HDP, n (%) |  |  |  |  | 0.28 |
| Gestational hypertension | 17 (49) | 10 (36) | 3 (27) | 2 (18) |  |
| Preeclampsia mild | 8 (23) | 6 (21) | 5 (45) | 2 (18) |  |
| Preeclampsia severe | 10 (29) | 12 (43) | 3 (27) | 7 (64) |  |
| Insurance, n (%) |  |  |  |  | 0.93 |
| No insurance | 1 (2.9) | 0 (0) | 0 (0) | 0 (0) |  |
| Private insurance | 26 (74) | 20 (71) | 7 (64) | 8 (73) |  |
| Public insurance | 8 (23) | 8 (29) | 4 (36) | 3 (27) |  |

<sup>†</sup> One-way analysis of means (not assuming equal variances); Kruskal-Wallis rank sum test; Fisher's exact test

Benjamini-Hochberg method to yield false discovery rates (FDR).^18^ Plots were generated using ggplot2, ComplexHeatmap, and Gmisc.^19–21^

## RESULTS

### Feasibility of home sample collection

In total, 129 individuals underwent two research visits through the Heart Health 4 New Moms study with 8 (6.2%) individuals completing the 2^nd^ visit in person prior to the COVID-19 pandemic and the remaining 121 (93.8%) individuals completing the visit remotely (**Supplemental Figure 1**). Of these 121 individuals who had remote study visits, 21 (17.4%) declined to provide a sample while 100 (82.6%) expressed willingness to complete the remote blood sample collection with the at-home self-collection kit. Of the 100 who agreed to sample collection, 89 participants (74% of the 121 eligible participants) successfully completed home self-sample collection. Two samples were lost in the return mail yielding a total of 87 home samples successfully collected through the study. Blood pressure measurements from the second study visit were missing for one participant who successfully provided a home sample, prompting the exclusion of that participant from the analysis.

### Participant characteristics and obstetric outcomes

Of the 87 remote samples successfully collected, 85 (97.7%) samples yielded usable proteomics data. The demographics, obstetric outcomes, and insurance status of the participants corresponding to these 85 samples are shown in **Table 1**. Based on blood pressure measurements taken at the second remote study visit, 35 individuals were normotensive (41.2%), 28 had Stage 1 HTN (32.9%), and 22 had Stage 2 HTN (25.8%). Of the 22 patients with Stage 2 HTN, 11 (50%) were taking antihypertensive medications. While HDP broadly are associated with an increased risk of progression to chronic hypertension, the incidence of HDP subtype (gestational hypertension, preeclampsia without or with severe features) did not differ between BP groups.^1,22^

Both pre-pregnancy and postpartum BMI was higher in those who were hypertensive compared to those who were normotensive, consistent with the previously reported increased risk of progression to chronic hypertension for individuals with obesity.^2^ However, Bonferroni post hoc analysis of postpartum BMI revealed significant differences only between Normotensive vs. Stage 2 HTN (p < 0.001), Stage 1 HTN vs. Stage 2 HTN (p = 0.003) and Normotensive vs. Stage 2 HTN with antihypertensive medications (p = 0.017). These findings suggest that, in a study in which all participants had BMI ≥ 25 kg/m^2^, a further elevation in BMI is associated with development of Stage 2 HTN but not with Stage 1 HTN.

### Differential expression analysis reveals protein levels altered in uncontrolled Stage 2 hypertension

Proteomic analysis of remote blood samples identified 381 unique proteins. Initially, principal component analysis (PCA) was used to explore the data and examine possible group clustering (**Figure 1a**). No clustering was observed, suggesting that the proteomes are generally heterogenous across the three groups defined here. Differential expression (DE) analysis was first performed to find proteins DE in Stage 1 HTN or Stage 2 HTN. DE proteins were defined as those proteins with Benjamini-Hochberg adjusted p-value (false discovery rate, FDR) < 0.05.

**Figure 1:**
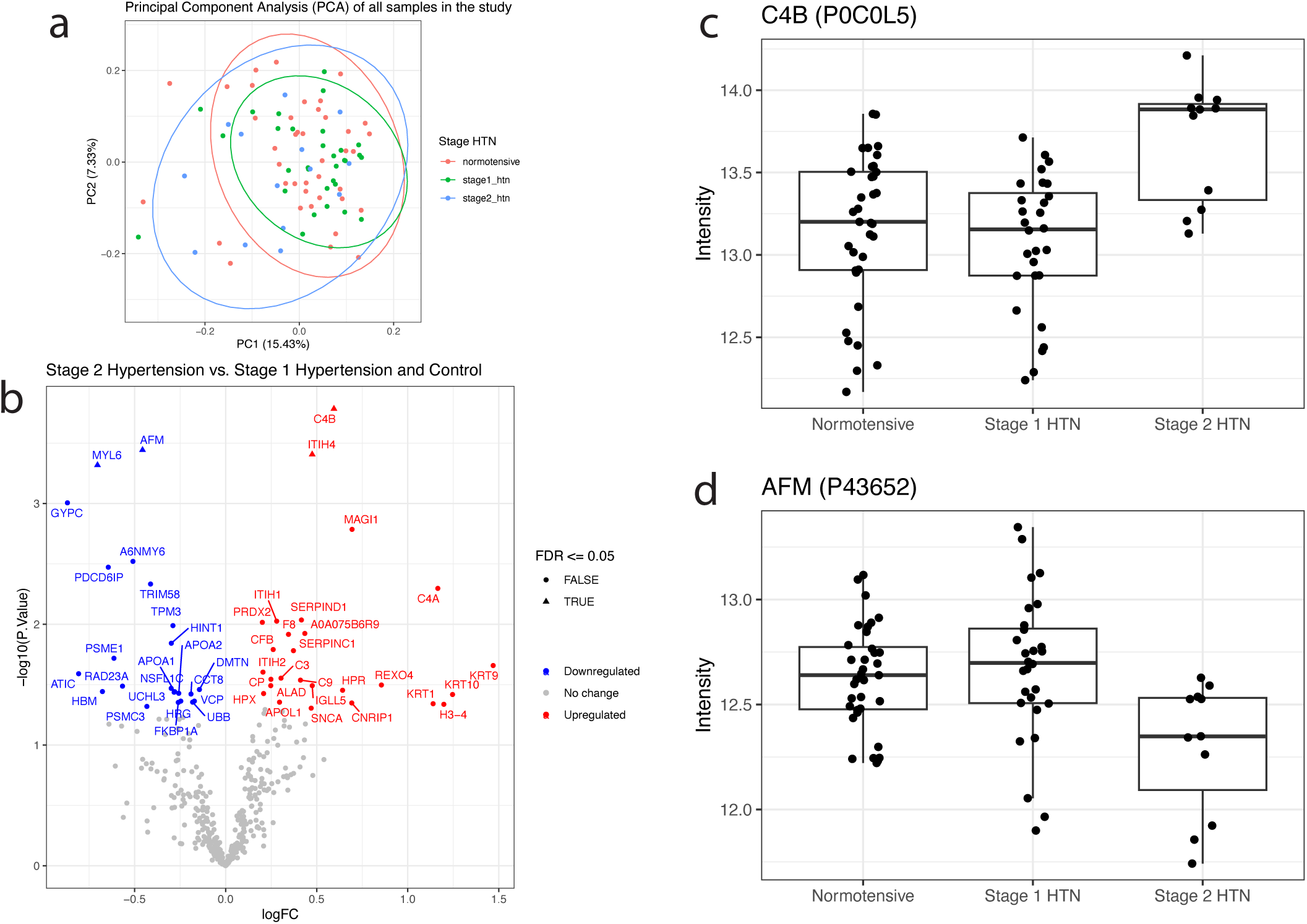
Differential expression analysis reveals protein levels altered in blood of patients with Stage 2 hypertension not taking antihypertensive mediciations. a) Principal component analysis of all 85 samples from which proteomic data was analyzed. b) Volcano plot of differential expression analysis results of individuals with Stage 2 hypertension not taking antihypertensive medications (n = 11) versus individuals with Stage 1 hypertension (n = 28) and normotension (n = 35). Proteins in red correspond to those upregulated in individuals with Stage 2 hypertension while those in blue are downregulated. Individual expression values for the most significantly downregulated (Afamin, AFM) and upregu-lated (Complement C4-B, C4B) are shown in c) and d). HTN = hypertension.

No proteins were DE in Stage 1 HTN (**Supplemental Table 1**), and one protein (Afamin, AFM) was significantly decreased in Stage 2 HTN (**Supplemental Table 2**). To explore the pathogenesis of uncontrolled postpartum HTN and avoid potential confounding effects of antihypertensive medications on serum protein levels (detailed above), we next disaggregated individuals with Stage 2 HTN based on whether they were taking antihypertensive medications. Comparing individuals with Stage 2 HTN who were not taking antihypertensive medications with those having Stage 1 HTN or normotension, and thus also not taking antihypertensive medications, revealed 4 DE proteins (**Figure 1b**). Complement C4-B (C4B, **Figure 1c**) and inter-alpha-trypsin inhibitor heavy chain 4 (ITIH4) were more abundant in blood of individuals

with Stage 2 hypertension relative to Stage 1 HTN or control (log_2_ fold change = 0.59 and 0.48, respectively), while myosin light chain 6 (MYL6) and Afamin (AFM, **Figure 1d**) were less abundant (log_2_ fold change = –0.70 and –0.45, respectively). Notably, there was high concordance between these results and those including all Stage 2 HTN patients, particularly with respect to AFM, which was DE in both, and C4B, which had the third smallest FDR in the first analysis and was DE in the second. To explore trends beyond these 4 differentially expressed proteins, we analyzed proteins with unadjusted p-value < 0.05. Hierarchical clustering on the 50 proteins which met this less stringent cutoff reveals partial separation between groups by heatmap visualization (**Supplemental Figure 2**).

### Functional analysis of differentially expressed proteins

We next performed Gene Ontology (GO) analysis of proteins DE in individuals with Stage 2 HTN not on antihypertensive medications. Given the low number of proteins which had FDR < 0.05 (n = 4), we performed GO analysis on those proteins which met the less stringent cutoff of unadjusted p-value < 0.05, allowing us to search for shared function among 27 and 23 proteins in the upregulated and downregulated searches, respectively (**Figure 2a**). The top GO term among upregulated proteins was “blood microparticle” (GO:0072562). DE proteins mapping to this term included the serine protease inhibitor SERPIN1C, complement component proteins C3, C4A, C4B, and C9, and inter-alpha-trypsin inhibitor heavy chains 1, 2, and 3 (**Supplemental Table 3**). Multiple GO terms relating to endopeptidase inhibition also showed enrichment of upregulated proteins, including endopeptidase inhibitor activity (GO:0004866), peptidase inhibitor activity (GO:0030414), and enzyme inhibitor activity (GO:0004857) (**Figure 2a**). Genes mapping to the endopeptidase inhibitor activity GO term include serine protease inhibitors SERPINC1 and SERPIND1 and early complement components C3, C4A, and C4B (**Supplemental Table 3**). The GO term “proteolysis involved in protein catabolic process” (GO:0051603) was the most significantly enriched among downregulated proteins (**Figure 2a**). Notable DE genes mapping to this GO term include proteasome subunits PSMC3 and PSME1,

**Figure 2:**
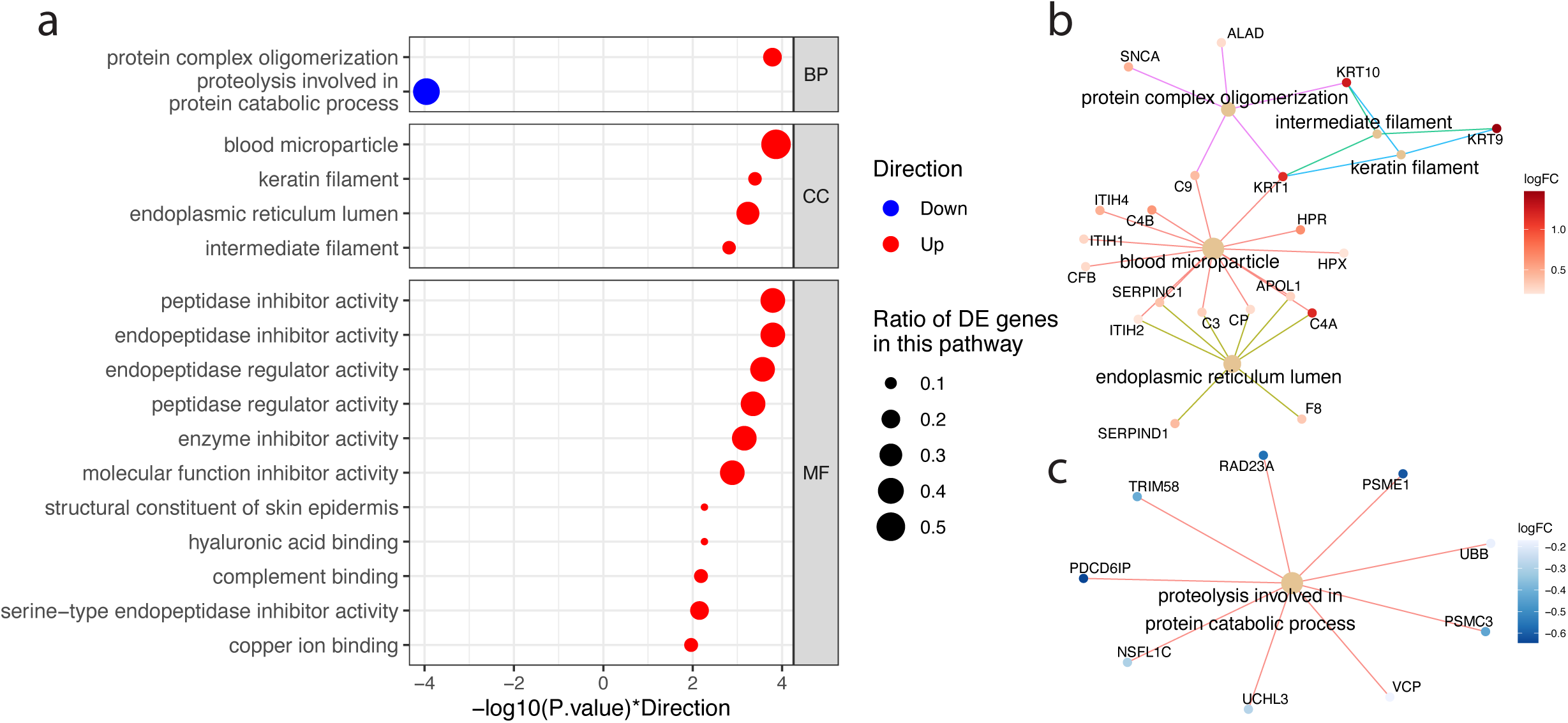
Functional analysis of proteins which are differentially expressed in individuals with Stage 2 hypertension not taking antihypertensive medications. a) GO terms which had p value < 0.01 for being enriched in proteins which were upregulated (red) or downregulated (blue) in serum of individuals with Stage 2 hypertension. These same results are plotted to show overlapping proteins for GO terms enriched in b) upregulated (red) and c) downregulated (blue) proteins. DE = differentially expressed.

UBB (encoding Ubiquitin B), and ubiquitin C-terminal hydrolase L3 (UCHL3) (**Supplemental Table 4**). All GO terms with p-value < 0.005 and the DE proteins mapping to them are shown in **Supplemental Table 3** and **Supplemental Table 4**. To visualize the complex and overlapping associations between genes and GO terms, we plotted gene-concept networks of top enriched GO terms among upregulated (**Figure 2b**) and downregulated (**Figure 2c**) proteins.

### Analysis of blood pressure changes across visits revealed unique markers of hypertension progression

As a secondary analysis, we sought correlations between the postpartum serum proteomics data at the second study visit and participant blood pressure trajectories over time. Blood pressure measurements from the two research study visits (at 8 and 46 weeks postpartum) were used to classify individuals into one of three groups as described above: improved HTN, worsened HTN, or no change in blood pressure (**Figure 3a**). Of the 35 individuals classified as “improved hypertension” by decreasing blood pressure, one started antihypertensive medications between the two study visits, while 3 discontinued medications. Similarly, of the 15 individuals classified as “worsened hypertension”, one initiated antihypertensive medications between these visits, while 4 discontinued medications (**Supplemental Table 5**).

**Figure 3:**
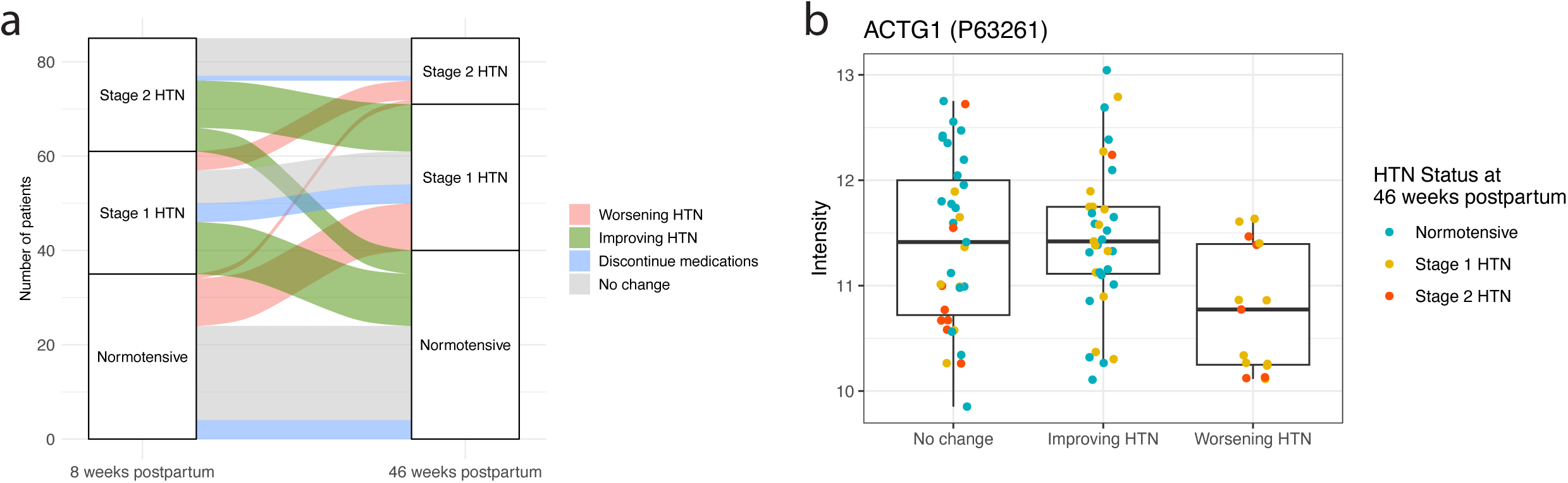
Blood pressure trajectory analysis reveals unique markers of chronic hypertension progression. a) Flow diagram depicting how participant blood pressure status changed from 8– to 46-weeks postpartum. b) Differential expression analysis revealed actin gamma 1 (ACTG1) as the most downregulated protein in serum of individuals with worsening HTN (n = 15) relative to improving HTN (n = 35) and unchanged HTN (n = 35, FDR = 0.054). HTN = hypertension.

We next performed differential expression analysis by comparing participants with worsened hypertension with those with unchanged or improved blood pressure. We discovered one protein, actin gamma 1 (encoded by ACTG1), which trended towards being downregulated in patients with worsened HTN (FDR = 0.054, **Figure 3b**). No other proteins had FDR < 0.10, though platelet basic protein, Filamin-A, Leucine-rich repeat-containing protein 53, and Cyclin-K were all downregulated in individuals with worsened HTN with unadjusted p-value < 0.01 (**Supplemental Table 6**). Notably, none of these proteins had unadjusted p-value < 0.01 in the primary analysis comparing HTN stages at the second study visit, suggesting that participants with worsened hypertension have unique proteomic signatures from those with Stage 2 hypertension.

## COMMENT

### Principal findings

In this study, we report a serum proteomic analysis of persistent hypertension following HDP collected through remote sampling devices. First, we demonstrate that postpartum individuals are willing and able to collect blood samples at home with 74% of those eligible successfully completing sample collection. Second, analysis of 381 serum proteins at one-year postpartum identified 4 proteins that were differentially expressed in individuals with new-onset Stage 2 hypertension following HDP and not taking antihypertensive medications (**Figure 1**). Functional enrichment analysis revealed a significant increase in proteins annotated to *blood microparticle* and *endopeptidase inhibitor-related* GO terms in blood of individuals with Stage 2 hypertension, and a decrease in proteins relating to *ubiquitin-proteasome mediated proteolysis* (**Figure 2**). A secondary analysis leveraging blood pressure patterns across the first-year postpartum uncovered changes in different proteins than the primary analysis, though none had FDR < 0.05 (**Figure 3**).

### Results in the context of what is known

While an association between HDP and later-life maternal CVD is well established, the underlying mechanisms are not well understood. Given the overlap of risk factors for cardiovascular disease and preeclampsia, some hypothesize that pregnancy is analogous to a cardiac stress test and that development of HDP simply represents a “failure” of this stress test.^5,23,24^ In the immediate postpartum period, however, those with HDP have significant cardiovascular dysfunction; whether these changes persist and are underlying drivers of long-term CVD is unclear.^25–27^ Persistent hypertension at one year postpartum in particular is associated with severe preeclampsia, suggesting that this facet of cardiovascular dysfunction may link HDP and CVD.^1^ Better understanding the biological pathways leading to chronic hypertension may be crucial to predicting and intervening to reduce CVD in patients who had HDPs. Together, the results reported in our analyses shed light on potential biological pathways leading to persistent hypertension following HDP.

### Clinical implications

The results of this study provide novel insight into persistent HTN after HDP and thus might inform future studies or yield novel drug targets. For example, our results suggest that inhibition of endopeptidases (e.g. neprilysin) and decreased activity of the ubiquitin-proteasome system may be involved in this disease process. Targeting these pathways pharmacologically, possibly with existing agents such as the neprilysin inhibitor sacubitril, may lead to novel clinical management strategies for persistent postpartum HTN.

### Research implications

We first show that 74% of eligible individuals successfully completed home blood sample collection. These findings indicate that this research methodology may be particularly appealing to this population with competing priorities in the postpartum period.^28^ From these samples, we conducted differential expression analyses to interrogate the pathogenesis of hypertension in those with prior HDPs. Four proteins were differentially expressed in blood of individuals with Stage 2 hypertension who were not taking antihypertensive medications. Complement C4-B (C4B), the most significantly upregulated protein, is part of the classic coagulation pathway C3 convertase.^29^ Coagulation cascade activation is a feature of many HDPs, and these findings suggest that its persistence may contribute to postpartum hypertension development.^5^ Inter-alpha-trypsin inhibitor heavy chain 4 (ITIH4), a protein that inhibits proteases such as endopeptidases, was also significantly upregulated.^30^ Among the downregulated proteins is MYL6, a component of the myosin light chain that has not been reported as a biomarker of any disease to our knowledge. mRNA expression of MYL6 increases in human vascular smooth muscle cells treated with preeclamptic plasma, however, possibly suggesting a causal relationship.^31^ The other significantly downregulated protein, Afamin (AFM), is a serum transport protein in the albumin gene family which is an early predictor of preeclampsia.^32,33^ This may further suggest that persistence of preeclamptic sequalae contributes to postpartum hypertension development.

Multiple GO terms relating to endopeptidase inhibition were enriched among upregulated proteins. Endopeptidases inactivate small peptides by cleaving them at specific, non-terminal amino acids sequences.^34^ For example, neprilysin is an endopeptidase that degrades natriuretic peptides, angiotensin II, and bradykinin.^35^ Neprilysin levels are elevated in preeclampsia, and dysregulation of natriuretic peptides is associated with HDP and development of hypertension at 2-7 years postpartum.^36–38^ While inhibition of neprilysin alone with sacubitril does not decrease blood pressure, combination therapy with valsartan is an effective therapy for hypertension.^39,40^ Our results suggest that dysregulation of endopeptidases might be implicated in hypertension development following HDPs.

Only one GO term, proteolysis involved in protein catabolic process, was significantly enriched among proteins downregulated in Stage 2 HTN. Many of these downregulated proteins are directly involved in ubiquitin-proteasome mediated proteolysis. Preeclampsia is associated with protein misfolding and elevated plasma proteasome levels.^41,42^ Our observation that ubiquitin-proteasome proteins are decreased in postpartum Stage 2 HTN might suggest that decreases in misfolded protein degradation contributes to hypertension progression. Furthermore, a recent study exploring serum proteomic profiles in early pregnancy showed matrix metalloproteinase 12 (MMP12), a protease, to be inversely associated with HDP development.^43^

While individuals with preeclampsia with severe features are at higher risk of developing hypertension in the first year postpartum, more granular characterizations of blood pressure recovery in this time period are needed.^1^ Inherent challenges to studying individuals in the postpartum period likely contribute to the lack of knowledge in this area.^6^ Our study helps address this gap in the literature by reporting matched blood pressure data from study visits at two time points. (**Figure 3, Supplemental Tables 5 and 6**). From this data, we defined 4 groups based on blood pressure changes across visits which correlate with different serum proteins than the blood pressure classifications at one year postpartum alone.

### Strengths and limitations

This study has strengths and weaknesses which must be considered. Remote in-home sample collection and study visits increased accessibility of research to a diverse cohort of postpartum individuals to overcome many of the barriers to engagement in research in the postpartum period. The proteins identified here require cross-validation, however, as this was a single-cohort study across two sites in one state. Further limiting generalizability, this study included only individuals with BMI ≥25 kg/m^2^ who had access to a device with internet. The disaggregation of patients with Stage 2 HTN based on whether they were taking antihypertensive medications could also introduce bias as there may be unaccounted for reasons that these patients are not receiving medical management for their hypertension. While the proteins dysregulated in new-onset postpartum Stage 2 HTN may reflect disease pathogenesis, they might instead simply reflect the effects of this disease process.

### Conclusions

In conclusion, we utilized serum proteomic analysis generated from remote blood sample collection to discover 4 proteins which are differentially expressed in individuals not taking antihypertensive medications who developed Stage 2 HTN following HDPs. Pathway enrichment analysis suggests that upregulated proteins have shared functions relating to endopeptidase inhibition, while downregulated proteins are involved in ubiquitin-proteasome mediated proteolysis. Stratifying patients by blood pressure changes across the first year postpartum reveals a distinct serum proteomic signature from this primary analysis. Together, these results suggest that protein homeostasis may be dysregulated in hypertension following HDPs and provide novel candidates for confirmation in larger cohorts and may serve as treatment targets for clinical interventions.

## Data Availability

All data produced in the present study are available upon reasonable request to the authors

## ACKNOWLEDGEMENTS

The authors are grateful for, and inspired by, the participants who devoted their time to the HH4NM study and furthermore provided remote blood samples all while caring for a newborn child during a global pandemic.

---

**Supplemental Figure 1:**
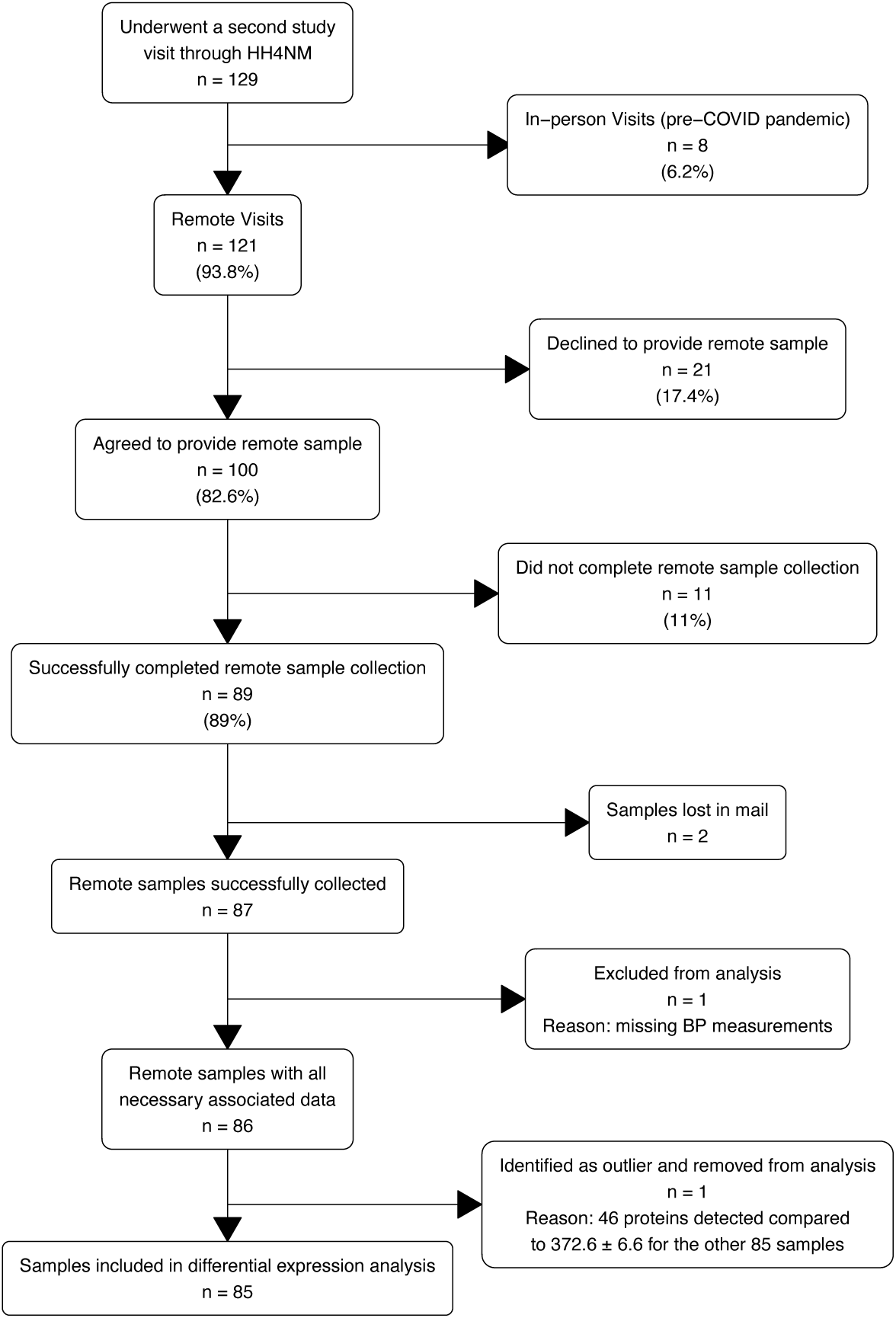
Enrollment flow diagram of home sample collection.

**Supplemental Figure 2:**
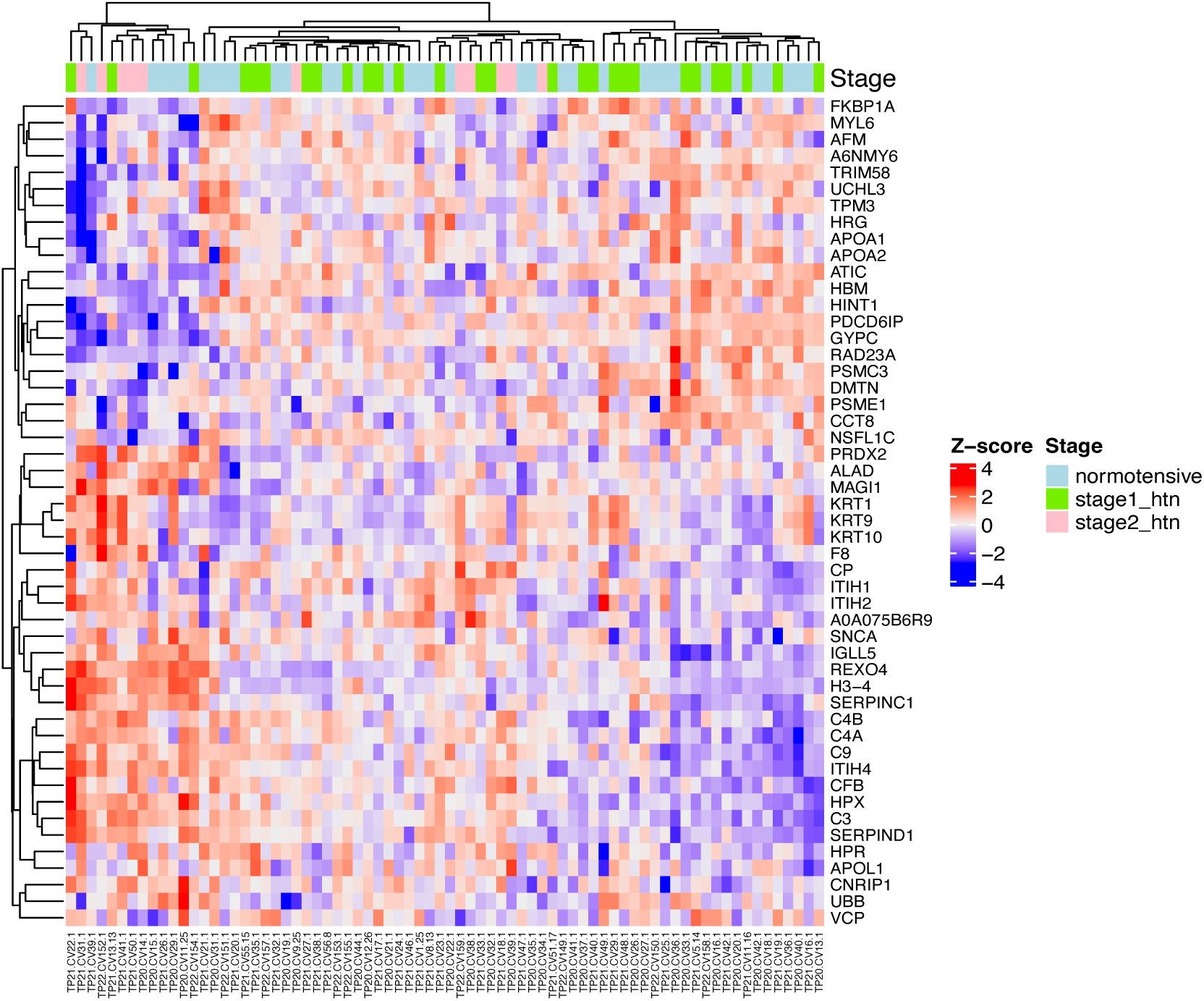
Heatmap of all proteins with unadjusted p value < 0.05 in individuals with Stage 2 HTN relative to Stage 1 HTN and normotension. Rows and columns were clustered using unsupervised hierarchical clustering.

**Supplemental Table 1:** Results of differential expression analysis comparing Stage 1 hypertension vs. Stage 2 hypertension and normotension. All proteins with unadjusted p-value < 0.05 are shown.

|  | logFC | AveExpr | t | P.Value | adj.P.Val | B | symbol |
| --- | --- | --- | --- | --- | --- | --- | --- |
| *P21980* | 0.3031829 | 12.40676 | 3.019345 | 0.003386391 | 0.8613492 | −2.767729 | TGM2 |
| *P43652* | 0.2384734 | 12.55750 | 2.821313 | 0.006012588 | 0.8613492 | −3.026523 | AFM |
| *P0C0L5* | −0.2878520 | 13.21640 | −2.697376 | 0.008499439 | 0.8613492 | −3.181961 | C4B |
| *P27348* | −0.4447394 | 11.64431 | −2.671178 | 0.009132595 | 0.8613492 | −3.214150 | YWHAQ |
| *P01593* | −0.4256019 | 13.48243 | −2.466076 | 0.015769716 | 0.8613492 | −3.457775 | IGKV1D−33 |
| *P48643* | −0.2879559 | 11.26761 | −2.329507 | 0.022321303 | 0.8613492 | −3.611452 | CCT5 |
| *Q16695* | −0.9398896 | 11.90381 | −2.326157 | 0.022508632 | 0.8613492 | −3.615133 | H3−4 |
| *Q96QZ7* | −0.3273134 | 15.63593 | −2.153524 | 0.034248497 | 0.8613492 | −3.798911 | MAGI1 |
| *Q5T3U5* | 0.4279615 | 12.56062 | 2.137620 | 0.035560198 | 0.8613492 | −3.815249 | ABCC10 |
| *Q8WUM4* | 0.3106214 | 12.74154 | 2.096435 | 0.039161828 | 0.8613492 | −3.857083 | PDCD6IP |
| *P01619* | −0.2021530 | 12.95485 | −2.075138 | 0.041144977 | 0.8613492 | −3.878445 | IGKV3−20 |
| *P02749* | 0.2336596 | 13.08292 | 2.054258 | 0.043172933 | 0.8613492 | −3.899210 | APOH |
| *Q9NZD4* | 0.4360805 | 12.70331 | 2.048089 | 0.043788336 | 0.8613492 | −3.905311 | AHSP |
| *Q9HC38* | −0.1524560 | 12.60296 | −2.047769 | 0.043820414 | 0.8613492 | −3.905627 | GLOD4 |
| *P49773* | 0.1679704 | 13.13641 | 2.023709 | 0.046294684 | 0.8613492 | −3.929268 | HINT1 |

**Supplemental Table 2:** Results of differential expression analysis comparing Stage 2 hypertension (including individuals taking antihypertensive medications) vs. Stage 1 hypertension and normotension. All proteins with unadjusted p-value < 0.05 are shown.

|  | logFC | AveExpr | t | P.Value | adj.P.Val | B | symbol |
| --- | --- | --- | --- | --- | --- | --- | --- |
| *P43652* | -0.4361017 | 12.55750 | -4.733346 | 9.238209e-06 | 0.003519758 | 3.1502990 | AFM |
| *P06753* | -0.2791199 | 13.76551 | -3.414912 | 9.990956e-04 | 0.160983108 | -0.8172178 | TPM3 |
| *P0C0L5* | 0.3885291 | 13.21640 | 3.340143 | 1.267584e-03 | 0.160983108 | -1.0162162 | C4B |
| *P0C0L4* | 0.9845519 | 12.29201 | 3.234443 | 1.764531e-03 | 0.168071557 | -1.2920093 | C4A |
| *P04921* | -0.5980491 | 12.36501 | -3.074756 | 2.870849e-03 | 0.194858461 | -1.6960048 | GYPC |
| *Q8WUM4* | -0.4929851 | 12.74154 | -3.052481 | 3.068637e-03 | 0.194858461 | -1.7511167 | PDCD6IP |
| *O75347* | -0.2990014 | 12.20624 | -2.996178 | 3.626373e-03 | 0.197378326 | -1.8890366 | TBCA |
| *P05546* | 0.3344897 | 13.35718 | 2.824594 | 5.956915e-03 | 0.253950438 | -2.2968808 | SERPIND1 |
| *P0DP23* | -0.4156673 | 12.10062 | -2.815168 | 6.118144e-03 | 0.253950438 | -2.3187306 | CALM1 |
| *Q96QZ7* | 0.4557297 | 15.63593 | 2.750821 | 7.330134e-03 | 0.253950438 | -2.4663178 | MAGI1 |
| *P60660* | -0.4075679 | 10.71364 | -2.750734 | 7.331902e-03 | 0.253950438 | -2.4665145 | MYL6 |
| *P22234* | -0.5338252 | 10.97081 | -2.648195 | 9.723014e-03 | 0.298577131 | -2.6959486 | PAICS |
| *P21980* | -0.2879658 | 12.40676 | -2.630982 | 1.018767e-02 | 0.298577131 | -2.7337606 | TGM2 |
| *P35527* | 1.2224119 | 10.02793 | 2.582409 | 1.160947e-02 | 0.298941913 | -2.8393625 | KRT9 |
| *A0A075B6R9* | 0.3257782 | 11.59305 | 2.547739 | 1.273149e-02 | 0.298941913 | -2.9137372 | IGKV2D-24 |
| *Q9NZD4* | -0.5882980 | 12.70331 | -2.534829 | 1.317367e-02 | 0.298941913 | -2.9412173 | AHSP |
| *P22314* | -0.2524937 | 11.77874 | -2.530113 | 1.333862e-02 | 0.298941913 | -2.9512266 | UBA1 |
| *P00450* | 0.2101313 | 12.76865 | 2.463779 | 1.586387e-02 | 0.335785153 | -3.0903509 | CP |
| *P00751* | 0.1954750 | 13.13669 | 2.429201 | 1.734322e-02 | 0.341037007 | -3.1616320 | CFB |
| *P11166* | 0.1897095 | 13.35869 | 2.361730 | 2.058886e-02 | 0.341037007 | -3.2982489 | SLC2A1 |
| *P01008* | 0.2733973 | 13.39428 | 2.345734 | 2.143337e-02 | 0.341037007 | -3.3301554 | SERPINC1 |
| *P35612* | 0.1591323 | 12.25942 | 2.343870 | 2.153373e-02 | 0.341037007 | -3.3338607 | ADD2 |
| *P02654* | -0.2514594 | 12.43763 | -2.321423 | 2.277567e-02 | 0.341037007 | -3.3782881 | APOC1 |
| *P02652* | -0.2116484 | 14.96461 | -2.312042 | 2.331320e-02 | 0.341037007 | -3.3967459 | APOA2 |
| *Q9UNZ2* | -0.2433459 | 11.00991 | -2.299851 | 2.402855e-02 | 0.341037007 | -3.4206378 | NSFL1C |
| *Q14624* | 0.2325223 | 13.13270 | 2.298202 | 2.412676e-02 | 0.341037007 | -3.4238605 | ITIH4 |
| *P63000* | 0.2509724 | 11.26713 | 2.297512 | 2.416798e-02 | 0.341037007 | -3.4252088 | RAC1 |
| *P27105* | 0.1652554 | 13.80092 | 2.259555 | 2.653265e-02 | 0.360561459 | -3.4988254 | STOM |
| *P06702* | -0.3423533 | 11.73758 | -2.242135 | 2.768445e-02 | 0.360561459 | -3.5322545 | S100A9 |
| *P01024* | 0.2280781 | 13.84225 | 2.223445 | 2.896890e-02 | 0.360561459 | -3.5678714 | C3 |
| *A6NMY6* | -0.2849360 | 13.97368 | -2.218222 | 2.933702e-02 | 0.360561459 | -3.5777772 | ANXA2P2 |
| *P32119* | 0.1247603 | 14.20439 | 2.153532 | 3.424779e-02 | 0.407762786 | -3.6987807 | PRDX2 |
| *P02549* | -0.1289148 | 12.60228 | -2.122380 | 3.685790e-02 | 0.425541204 | -3.7559267 | SPTA1 |
| *O43707* | -0.3693022 | 12.60290 | -2.074793 | 4.117784e-02 | 0.453492611 | -3.8417989 | ACTN4 |
| *B9A064* | 0.3349700 | 14.28163 | 2.035331 | 4.508490e-02 | 0.453492611 | -3.9116948 | IGLL5 |
| *P02647* | -0.2018368 | 13.84572 | -2.028431 | 4.579984e-02 | 0.453492611 | -3.9237939 | APOA1 |
| *P13645* | 0.9035973 | 10.25614 | 2.023827 | 4.628223e-02 | 0.453492611 | -3.9318460 | KRT10 |
| *P04264* | 0.8427879 | 11.97798 | 1.993354 | 4.958633e-02 | 0.453492611 | -3.9847296 | KRT1 |

**Supplemental Table 3.** All individual proteins mapping to GO terms with p value < 0.005 which were enriched in upregulated proteins in serum of individuals with Stage 2 HTN relative to Stage 1 HTN and normotension.

|  | Term | Ont | N | DE | P.DE | mapped_genes |
| --- | --- | --- | --- | --- | --- | --- |
| GO:0072562 | blood microparticle | CC | 72 | 14 | 0.0001320442 | SERPINC1, CFB, C3, C4A, C4B, C9, CP, HPR, HPX, ITIH1, ITIH2, ITIH4, KRT1, APOL1 |
| GO:0051259 | protein complex oligomerization | BP | 9 | 5 | 0.0001818616 | ALAD, C9, KRT1, KRT10, SNCA |
| GO:0045095 | keratin filament | CC | 3 | 3 | 0.0004004325 | KRT1, KRT9, KRT10 |
| GO:0043588 | skin development | BP | 3 | 3 | 0.0004004325 | KRT1, KRT9, KRT10 |
| GO:0005788 | endoplasmic reticulum lumen | CC | 29 | 8 | 0.0005703659 | SERPINC1, C3, C4A, CP, F8, SERPIND1, ITIH2, APOL1 |
| GO:0004866 | endopeptidase inhibitor activity | MF | 33 | 8 | 0.0015001446 | SERPINC1, C3, C4A, C4B, SERPIND1, ITIH1, ITIH2, ITIH4 |
| GO:0030414 | peptidase inhibitor activity | MF | 33 | 8 | 0.0015001446 | SERPINC1, C3, C4A, C4B, SERPIND1, ITIH1, ITIH2, ITIH4 |
| GO:0004857 | enzyme inhibitor activity | MF | 41 | 9 | 0.0015155771 | SERPINC1, C3, C4A, C4B, SERPIND1, ITIH1, ITIH2, ITIH4, SNCA |
| GO:0005882 | intermediate filament | CC | 4 | 3 | 0.0015197422 | KRT1, KRT9, KRT10 |
| GO:0045109 | intermediate filament organization | BP | 4 | 3 | 0.0015197422 | KRT1, KRT9, KRT10 |
| GO:2000427 | positive regulation of apoptotic cell clearance | BP | 4 | 3 | 0.0015197422 | C3, C4A, C4B |
| GO:0002252 | immune effector process | BP | 43 | 9 | 0.0022032415 | CFB, C3, C4A, C4B, C9, HPX, KRT1, PRDX2, IGLL5 |
| GO:0061135 | endopeptidase regulator activity | MF | 35 | 8 | 0.0022916378 | SERPINC1, C3, C4A, C4B, SERPIND1, ITIH1, ITIH2, ITIH4 |
| GO:0140678 | molecular function inhibitor activity | MF | 44 | 9 | 0.0026318061 | SERPINC1, C3, C4A, C4B, SERPIND1, ITIH1, ITIH2, ITIH4, SNCA |
| GO:0061134 | peptidase regulator activity | MF | 37 | 8 | 0.0033862791 | SERPINC1, C3, C4A, C4B, SERPIND1, ITIH1, ITIH2, ITIH4 |
| GO:0019724 | B cell mediated immunity | BP | 22 | 6 | 0.0035695333 | C3, C4A, C4B, C9, HPX, IGLL5 |
| GO:0016064 | immunoglobulin mediated immune response | BP | 22 | 6 | 0.0035695333 | C3, C4A, C4B, C9, HPX, IGLL5 |
| GO:0008544 | epidermis development | BP | 5 | 3 | 0.0036051229 | KRT1, KRT9, KRT10 |
| GO:0030212 | hyaluronan metabolic process | BP | 5 | 3 | 0.0036051229 | ITIH1, ITIH2, ITIH4 |
| GO:0045104 | intermediate filament cytoskeleton organization | BP | 5 | 3 | 0.0036051229 | KRT1, KRT9, KRT10 |
| GO:0045103 | intermediate filament-based process | BP | 5 | 3 | 0.0036051229 | KRT1, KRT9, KRT10 |
| GO:1903510 | mucopolysaccharide metabolic process | BP | 5 | 3 | 0.0036051229 | ITIH1, ITIH2, ITIH4 |
| GO:0051262 | protein tetramerization | BP | 5 | 3 | 0.0036051229 | KRT1, KRT10, SNCA |
| GO:2000425 | regulation of apoptotic cell clearance | BP | 5 | 3 | 0.0036051229 | C3, C4A, C4B |
| GO:0002460 | adaptive immune response based on somatic recombination of immune receptors built from immunoglobulin superfamily domains | BP | 23 | 6 | 0.0045761275 | C3, C4A, C4B, C9, HPX, IGLL5 |

**Supplemental Table 4.**
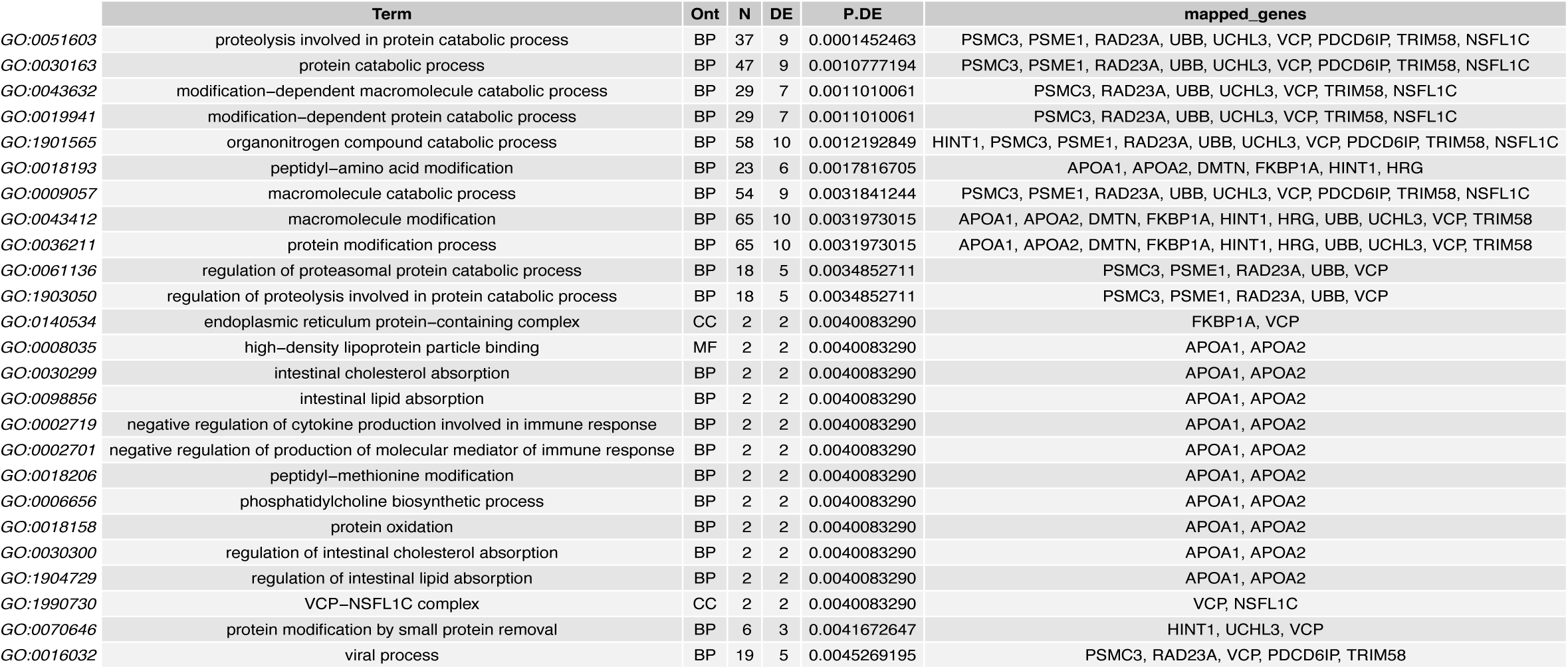
All individual proteins mapping to GO terms with p value < 0.005 which were enriched in downregulated proteins in serum of individuals with Stage 2 HTN relative to Stage 1 HTN and normotension.

**Supplemental Table 5.** Tabulated data depicting how many individuals were classified as normotensive or as having Stage 1 Hypertension or Stage 2 Hypertension at postpartum study visit 1 (8 weeks postpartum) and postpartum study visit 2 (46 weeks postpartum). “Change med status” represents whether individuals stopped or started antihypertensive medications between the two visits.

| visit1_stage_htn | visit2_stage_htn | bp_trajectory | change_med_status | n |
| --- | --- | --- | --- | --- |
| Normotensive | Normotensive | Improving HTN | change_meds | 4 |
| Normotensive | Normotensive | No change | no_change | 20 |
| Stage 1 HTN | Normotensive | Improving HTN | no_change | 11 |
| Stage 2 HTN | Normotensive | Improving HTN | no_change | 5 |
| Normotensive | Stage 1 HTN | Worsening HTN | no_change | 10 |
| Stage 1 HTN | Stage 1 HTN | Improving HTN | change_meds | 4 |
| Stage 1 HTN | Stage 1 HTN | No change | no_change | 7 |
| Stage 2 HTN | Stage 1 HTN | Improving HTN | no_change | 10 |
| Normotensive | Stage 2 HTN | Worsening HTN | no_change | 1 |
| Stage 1 HTN | Stage 2 HTN | Worsening HTN | no_change | 4 |
| Stage 2 HTN | Stage 2 HTN | Improving HTN | change_meds | 1 |
| Stage 2 HTN | Stage 2 HTN | No change | no_change | 8 |

**Supplemental Table 6.** Proteins which are differentially expressed with unadjusted p value < 0.01 in individuals with worsening postpartum hypertension relative to individuals with improving and unchanged hypertension.

|  | logFC | AveExpr | t | P.Value | adj.P.Val | B | symbol |
| --- | --- | --- | --- | --- | --- | --- | --- |
| *P63261* | −0.8640397 | 11.31362 | −3.992315 | 0.0001429615 | 0.05446834 | 0.3882896 | ACTG1 |
| *P02775* | −0.4188569 | 13.43115 | −3.079315 | 0.0028318778 | 0.53947272 | −1.8073917 | PPBP |
| *P21333* | −0.2391838 | 12.03555 | −2.785001 | 0.0066614575 | 0.66395885 | −2.4268727 | FLNA |
| *A6NM62* | −0.5535768 | 15.60813 | −2.737604 | 0.0076047904 | 0.66395885 | −2.5220983 | LRRC53 |
| *O75909* | −1.2627899 | 12.89133 | −2.688333 | 0.0087133707 | 0.66395885 | −2.6197112 | CCNK |

